# Estimated Spike Evolution and Impact of Emerging SARS-CoV-2 Variants

**DOI:** 10.1101/2021.05.06.21256705

**Authors:** Yong Lu, Kun Han, Gang Xue, Ningbo Zheng, Guangxu Jin

**Affiliations:** Department of Microbiology and Immunology, Wake Forest School of Medicine, Winston-Salem, NC 27101; Department of Cancer Biology, Wake Forest School of Medicine, Winston-Salem, NC 27157; Wake Forest Baptist Comprehensive Cancer Center, Winston-Salem, NC, USA; Center for Precision Medicine, Wake Forest School of Medicine, Winston-Salem, NC 27157

## Abstract

The severe acute respiratory syndrome coronavirus 2 (SARS-CoV-2), the virus that causes COVID-19, has been mutating and thus variants emerged. This suggests that SARS-CoV-2 could mutate at an unsteady pace. Supportive evidence comes from the accelerated evolution which was revealed by tracking mutation rates of the genomic location of Spike protein. This process is sponsored by a small portion of the virus population but not the largest viral clades. Moreover, it generally took one to six months for current variants that caused peaks of COVID-19 cases and deaths to survive selection pressure. Based on this statistic result and the above speedy Spike evolution, another upcoming peak would come around July 2021 and disastrously attack Africa, Asia, Europe, and North America. This is the prediction generated by a mathematical model on evolutionary spread. The reliability of this model and future trends out of it comes from the comprehensive consideration of factors mainly including mutation rate, selection course, and spreading speed. Notably, if the prophecy is true, then the new wave will be the first determined by accelerated Spike evolution.

## Introduction

According to the statistics of World Health Organization (https://covid19.who.int/), COVID-19 pandemic has caused at least 131 million confirmed cases and more than 2.8 million deaths as of April 4, 2021. During global circulating, the SARS-CoV-2 has been mutating despite at a slower rate than influenza and human immunodeficiency virus (HIV) (*1-5*). Not surprisingly, a notorious variant named B.1.1.7 has infected more people than other strains (*6*). Its occurrence not only raised new questions on the higher infectivity and fatality, but also suggested uneven evolution paces on variants’ mutation and selection, separately and uniquely. Thus, new bioinformatics tools are in urgent demand to develop to answer and confirm if a dynamic evolution could be true by tracking mutation rate.

To achieve this goal, efforts have been devoted to studying viral phylodynamic by finding and analyzing clades (*3*). GISAID (https://www.gisaid.org) maintains complete SARS-CoV-2 genomes and had more than 800,000 genomes in the database as of March 2021. The phylogenetic analysis has identified 9 clades in GISAID which are S, L, O, V, G, GH, GV, GR, and GRY. Among them, clades S and O were the most common ones in January and February 2020, whereas clades G and GRY prevailed from December 2020 to March 2021, especially the latter one. Given the widespread appearance and large numbers of B.1.1.7 genomes globally surpassing numbers of other existing clades, GISAID elevated the B.1.1.7 lineage to clade GRY that was descended from clade GR.

More importantly, these data have facilitated the mutation surveillance on Spike protein to understand how the pandemic shifts characteristics over time. A major form of the Spike mutations is D614G (*3, 7-10*). It reduces S1 shedding and increases infectivity. Also, it is the marker mutation for clades G, GH, GV, GR, and GRY.

Unexpectedly, at the genomic level, B.1.1.7 not only accumulated mutations that failed nucleic acid testing (*11*), but have more nucleotide changes on the genomic region of the C-terminal domain of Spike protein with yet unknown importance (*12*). Accordingly, took nonsense mutation together with all other types into consideration, we developed a bioinformatics model to study (1) the speed of Spike genomic evolution and further (2) how long it would take by a new variant to ignite another outburst. Based on and supported by this model, we found that most emerging variants were generated from a small portion of relatively original mutated strains driven by accelerated evolution. Furthermore, newly emerged variants cannot immediately cause a pandemic peak but need to show highly selective advantages within one to six months among a large population. These findings could benefit a scientific understanding of the dynamic virus evolution and benefit practical predicting of an upcoming wave.

## Results

### Mutation Rate of Spike Genomic Sequence

Our bioinformatics approach recorded mutation numbers of mismatches and indels in Spike coding region by first aligning GISAID-derived 817,351 SARS-CoV-2 sequences as of March 15^th^ 2020 to the WIV04-Spike reference (*13*), using the BLAST software (*5*) (**Methods**), and then normalized them to the length of the Spike genomic region. Next, Mutated Number per Kilobase (MNPK) was calculated by multiplying the normalization results by 1,000 and defined as Spike mutation rate (**Fig. S1**).

Further analysis showed that in gradually disappearing clades (L, O, S, and V), high MNPKs densely happened in early time points but later disappeared or dramatically decreased to a low-frequency level where dots were way sparse and values below 5, most around 2.5 (**Fig. 1**). While in currently prevalent clades (G, GH, GV, and GR), oppositely, high MNPKs clearly increased to a serious enrichment in at least one continent (**Fig. 1**). This suggested that these Spike coding regions are still swiftly mutating without an obvious peak thus new variants could continuously appear. Furthermore, steady highest MNPKs only showed at most recent time points in the newest clade GRY (*14*), especially in Europe and North America (**Fig. 1**). Lastly, SARS-CoV-2 genomes not assigned (NA) into any clades defined by GISAID also showed some mutations outnumbered by GRY but still accumulated in 3 continents (**Fig. 1**). A question that if another new variant would come out of them doesn’t have an answer yet but need long-term monitoring.

**Figure 1:**
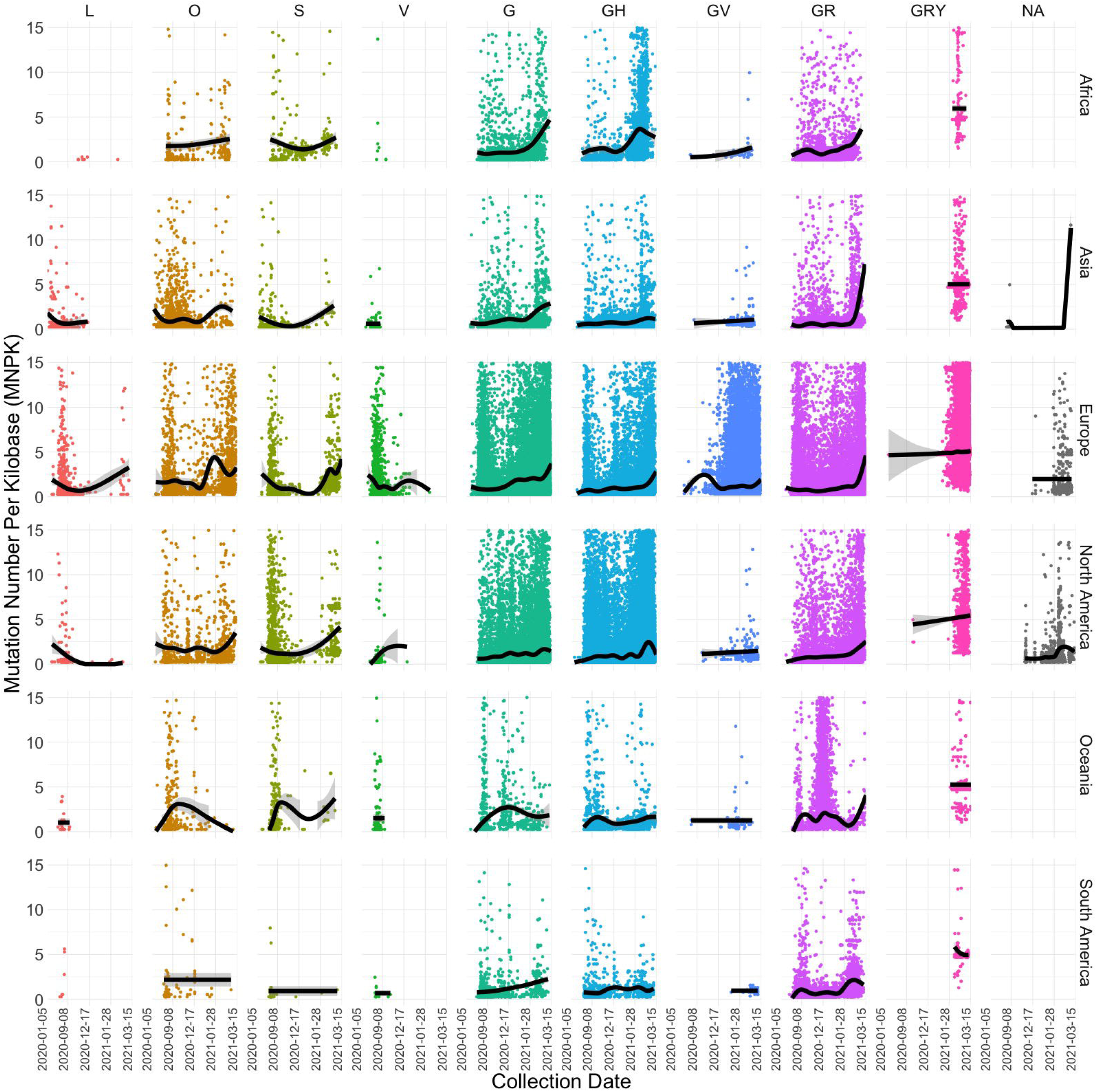
Mutation rate changes over the global pandemic. MNPK values (vertical ordinate) are illustrated by clade (column) and continent (row) from January 5^th^ 2020 to March 15^th^ 2021 (horizontal ordinate, segmented by every 100 sampling days). Each single dot represents a mutation rate of one SARS-CoV-2 sequence calculated after blasting. Columns are titled by CISAID clades and arranged from left to right according to their relative chronological order. Not assigned (NA) means SARS-CoV-2 sequences carrying mutations but are not classified into a new clade yet according to GISAID’s definition. Rows are titled by continents where SARS-CoV-2 sequences were collected and are arranged from top to bottom by alphabetical order. Black lines: fitting curves generated by Locally Estimated Scatterplot Smoothing (LOWESS). Gray shades: 95% confidence level interval.

Taken together, our bioinformatic approach detected a trend of Spike mutation rates which is in accordance with the relative chronological order of each clade’s occurrence yet not necessarily associated with the population size of continents.

### Relative Evolutionary Rate of Spike Genomic Sequence (RERS)

#### Defining Spike Evolutionary Rate

Since the trend we observed is quite similar to reported significantly increased mutation rates on emerging variants such as B.1.1.7 (*8, 9, 12, 14*), we then reasonably hypothesized that the differences in Spike genomic mutation rates could reflect a revolutionary strategy in which its pace was elevated for SARS-CoV-2 to generate more infectious variants, no matter from single mutation or recombination (*15*), and dropped after task completion then precedent variants disappeared. This process could be a result of accelerated evolution or viral recombination. If this is true, then Spike genomic region evolves at different speeds in different variants.

To verify the hypothesis, we defined **Relative Evolutionary Rate of Spike** (RERS) by MNPK over time (**Methods**) and measured it according to (i) to each clade in a continent, SARS-CoV-2 sequences were grouped every seven sampling days from January 2^nd^ 2020 to March 15^th^ 2021 and arithmetic mean MNPKs were calculated; (ii) RERS_*i*_ (*i* = 2, 3, 4, …, g) was calculated by the following equation.

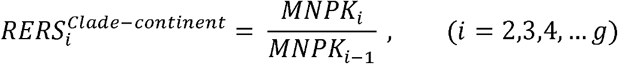

Finally, after deduction of 1 from 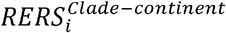, positive and negative values mean increased and decreased Spike evolution, respectively.

#### Spike Evolution Showed Differentiated Speeds over the Course of the Pandemic

Through further analysis at the clade and geographic-region level, it showed that to gradually disappearing clades (L, O, S, and V), 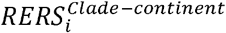 values either cannot be observed or sparse, suggesting that Spike genomic region in these four clades stopped or hugely slowed down their revolution, consistent with the fact that these clades are gradually disappearing from the pandemic. On the contrary, an obvious acceleration can be observed in currently prevalent clades G, GH, GV, and GR (**Fig. 2**). Besides, in each continent, at least one clade is evolving at an elevated speed, in Europe even all four. This clearly suggested that these clades are still globally dominant and undergoing speed-up evolution everywhere.

**Figure 2:**
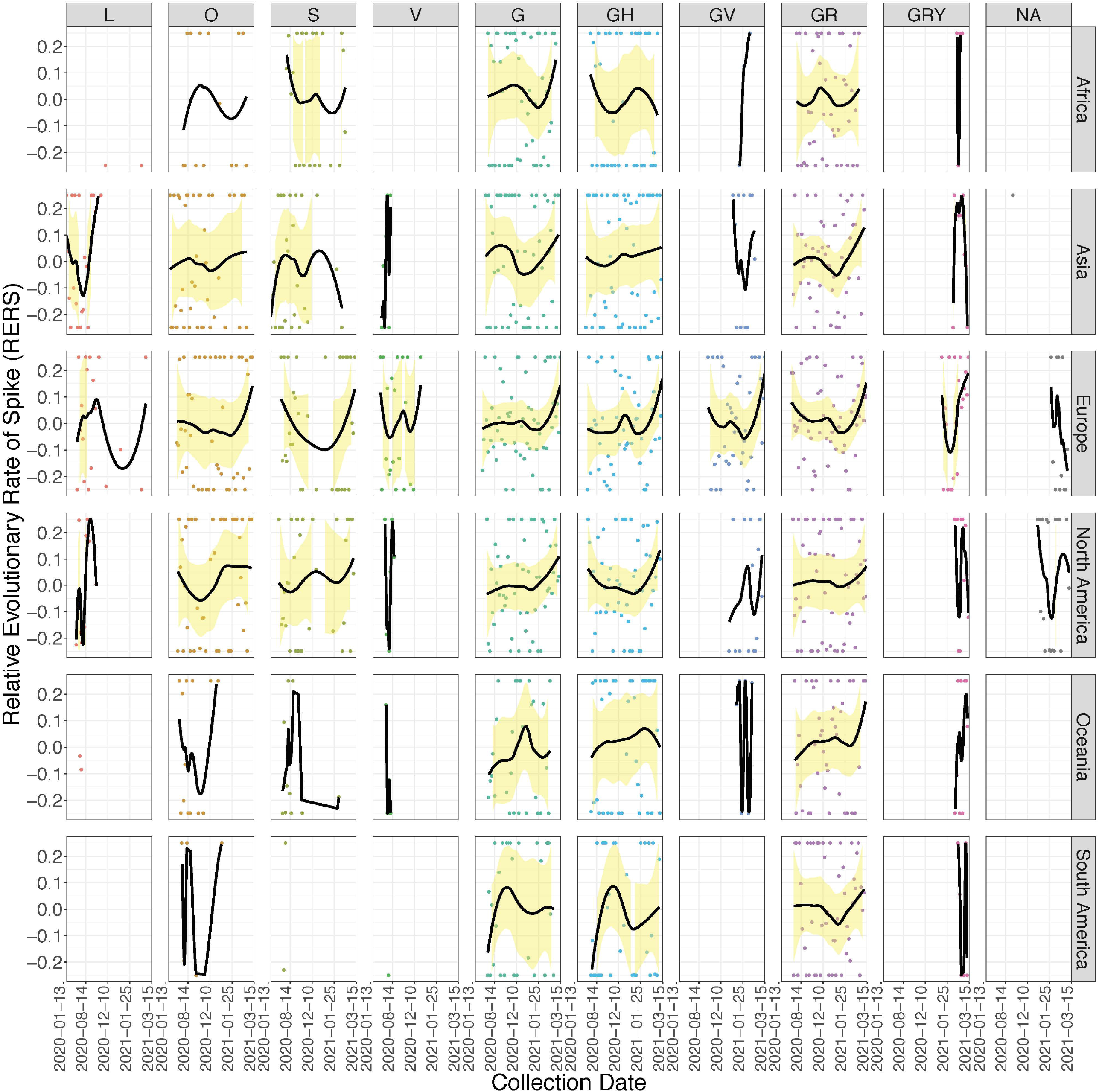
Evolutionary rate changes over the global pandemic. Evolutionary rates of Spike genomic region (vertical ordinate) are illustrated by clade (column) and continent (row) from January 13^th^ 2020 to March 15^th^ 2021 (horizontal ordinate). Each single dot represents an RERS value. Columns are titled by CISAID clades and arranged from left to right according to their relative chronological order. NA means SARS-CoV-2 sequences carrying mutations but have not been classified into a new clade yet according to GISAID’s definition. Rows are titled by continents where SARS-CoV-2 sequences were collected and are arranged from top to bottom by alphabetical order. Black lines: fitting curves generated by Gamma Regression. Yellow shades: 95% confidence level interval.

Next, 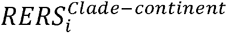 in clade GRY hasn’t increased since February 2021 (**Fig. 2**) though it has the highest MNPK (**Fig. 1**). This may be attributed to the huge selective pressure brought about by vaccines, together with the fact that not enough time nor input sequences specifically for this clade. So, the evolutionary situation of GRY should still be monitored.

Finally, one increased and one decreased RERS were observed from NA SARS-CoV-2 sequences in Europe and North America (**Fig. 2**) with currently unclear influence.

Conclusively, 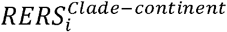 distribution showed that faster Spike evolution is driven by accelerated mutation in its genomic region thus governs the ongoing pandemic and/or potentially generates new variants possibly responsible for next wave, and thus confirmed our hypothesis.

#### A Global Perspective of Spike Evolution

We further explore more potentials of RERS in a more general manner. Similar to 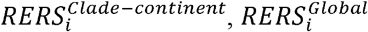 was calculated by all SARS-CoV-2 sequences but without restrictions from clades and continents.

From 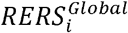 fitting curve, we can clearly see a three-staged, two-turning-point pattern (**Fig. 3A**): It initially vigorously rocketed from January 13^th^ 2020 to the peak on May 29^th^ 2020, then experienced a long-time slowly “decreasing” till reached the valley on October 26^th^ 2020, and restored a fast increasing by March 15^th^ 2021 without any sign of slowing down. We analyzed SARS-CoV-2 genomic diversity on the days of 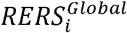 and COVID-19 peaks, one day randomly picked during the first rising phase and two more days in the second rising phase. We found that in the first rocketing stage, clades V and O were the most two abundant forms.

**Figure 3:**
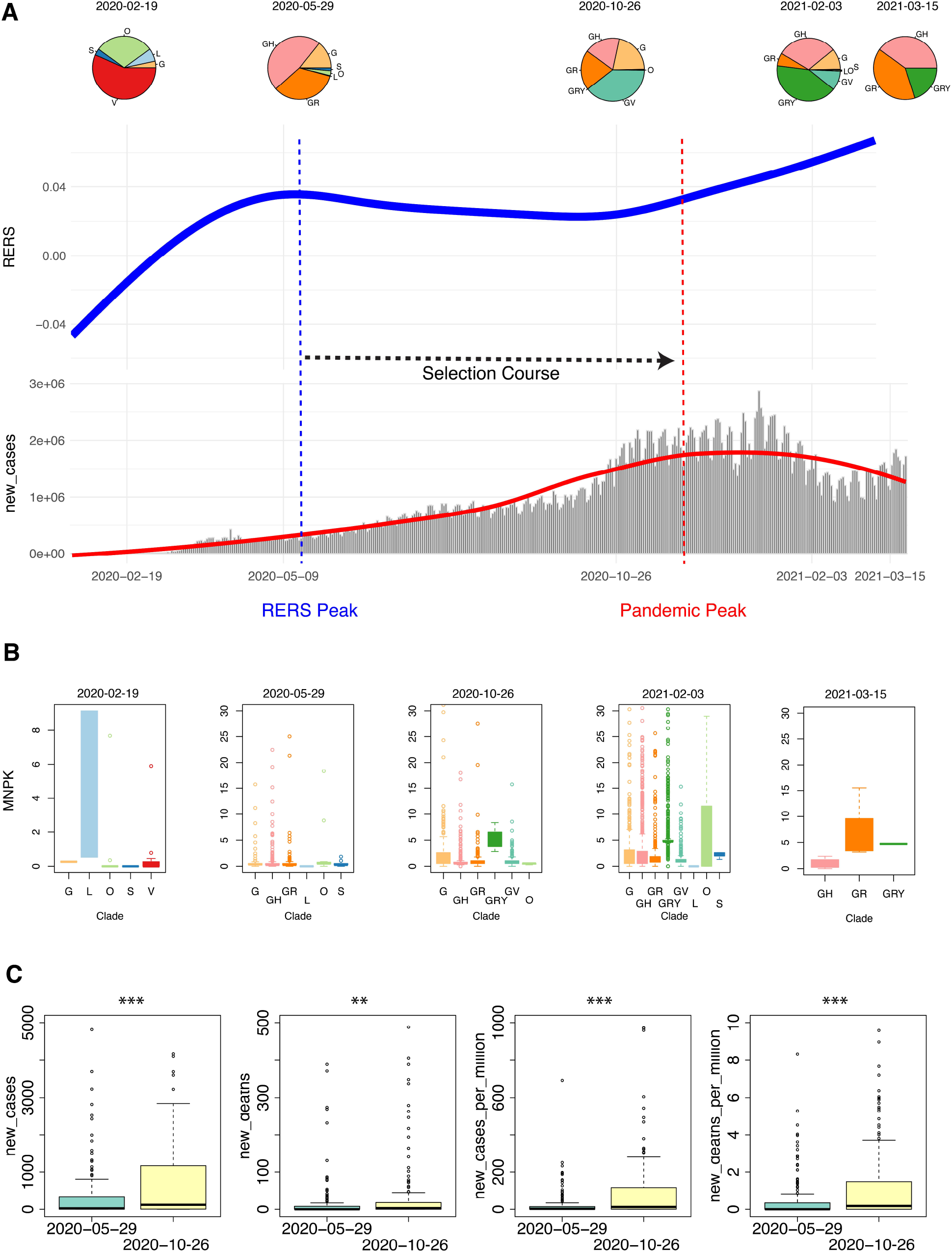
Global perspective of Spike evolution. (**A**) RERS shows a three-staged, two-turning-point pattern. A selection course is between RERS peak and pandemic peak. Blue line: RERS fitting curve generated using all SARS-CoV-2 sequence data in GISAID by March 15^th^ 2021. Red line: COVID-19 fitting curve using data of new cases by March 23^rd^ 2021. Gray bars: global new cases per day. Each pie chart shows SARS-CoV-2 genomic diversity on a representative sampling date. Fitting curves were generated by LOWESS. (**B**) MNPK values on each sampling day’s clades. Each single dot represents one outlier. (**C**) Numbers of the four pandemic parameters on May 29^th^ 2020 (cyan box) and December 26^th^ 2020 (yellow box). Each single dot represents one outlier. *P* values were calculated by Wilcoxon Rank-Sum Test. *: *P* < 0.05, **: *P* < 0.01, ***: *P* < 0.001.

Differently, at the peak, clades O, S, and L together shrank to a very small portion while clades G, GH, and GR gained dominance. This situation lasted to the valley where not only did clades (G, GH, GV, and GR) absolutely constituted almost all genomic forms but a new clade GRY developed. Then in the second increasing stage, clades GRY and GH obviously enlarged their genomic percentage until March 15^th^ 2021 when GRY ratio diminished but clades GH and GR were still strongly maintaining their dominant position (**Fig. 3A**). Thus, 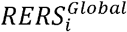 could be used to trace global Spike evolution trend.

Since variation of clades genomic distribution was observed at different stages of the 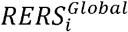 pattern, we further reasonably guessed if dominant clades could be the driving force for increased Spike evolution. To provide an answer, we turned back to compare MNPK of all clades at each selected time point. Quite differed from distribution results, clade L, but not the largest V, on February 19^th^ 2020 showed overwhelmingly high MNPK over the rest (**Fig. 3B**), suggesting that it was contributing the most mutations to the first rising phase, at least at that point. This characteristic ended on May 29^th^ 2020 when mutation and revolution all reached the peak (**Fig. 3B**). On October 26^th^ 2020 when the wave was about to start, most SARS-CoV-2 Spike mutations were generated from clade GRY. In the second climbing phase, clades O and GR had the most mutations (**Fig. 3B**). These data together suggested that during a rapid mutation-accumulating stage, the largest contribution doesn’t necessarily come from the biggest clades while might from a very small portion of viruses.

After fitting the 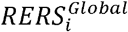 pattern, we then observed if it could be associated with COVID-19 data of new cases and deaths by fitting worldwide pandemic data (new cases per day) in the same image from WHO Coronavirus (COVID-19) Dashboard (https://covid19.who.int/) and Our World in Data (https://ourworldindata.org/) by March 23^rd^ 2021. Through comparison, there was a six-month course from 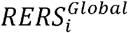 peak on May 29^th^ 2020 to that of the pandemic peak on November 28^th^ 2020 (**Fig. 3A**). We then compared new_cases, new_deaths, new_cases_per_million_people, and new_deaths_per_million_people of the two days to gain more insights into the pandemic and found that numbers of 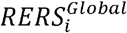 peak were significantly lower than those of the pandemic peak no matter in which comparisons (**Fig. 3C**). Our results suggested that it took time for finally accumulated Spike mutations to truly and eventually cause mass infection and death in population, like the reported one-month selection time for D614G (*3*) and matched the selection process in the evolutionary spread (*16, 17*). Such time was about six months in our bioinformatics approach.

Taken together, through the calculation of 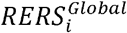 we showed a global Spike revolutionary pattern and a phenomenon that small portions of viruses might contribute more to mutations required for new variants. Further comparison between peaks of 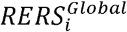 and global pandemic revealed a time course that possibly served as the selection stage for newly generated variants to cause next surge in COVID-19 cases and deaths.

#### Estimated Selection Course for All Continents

To estimate selection courses for all continents, we adopted 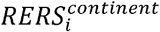 by pooling all clades including NA data. Similarly, we also observed that COVID-19 data peaks occurred after that of RERS in each continent. This suggested that the selection course we observed using global data also existed in all continents (**Fig. 4, Fig. S2**). Based on the high consistency between the collected SARS-CoV-2 sequence number and the new case number every day (**Fig. S3**), there is no system error caused by the data we used here. This on one hand showed a good match with the reported one-month (*3*) and our model-calculated six-month selection time (**Fig. 3A**), and on the other hand clearly illustrated that the length of selection could largely determine when a next pandemic peak could come. Thus, our results suggested that although 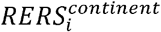 could not predict pandemic in a real-time manner, its most recent peaks could serve as an important indicator of the following waves of infections and deaths.

**Figure 4:**
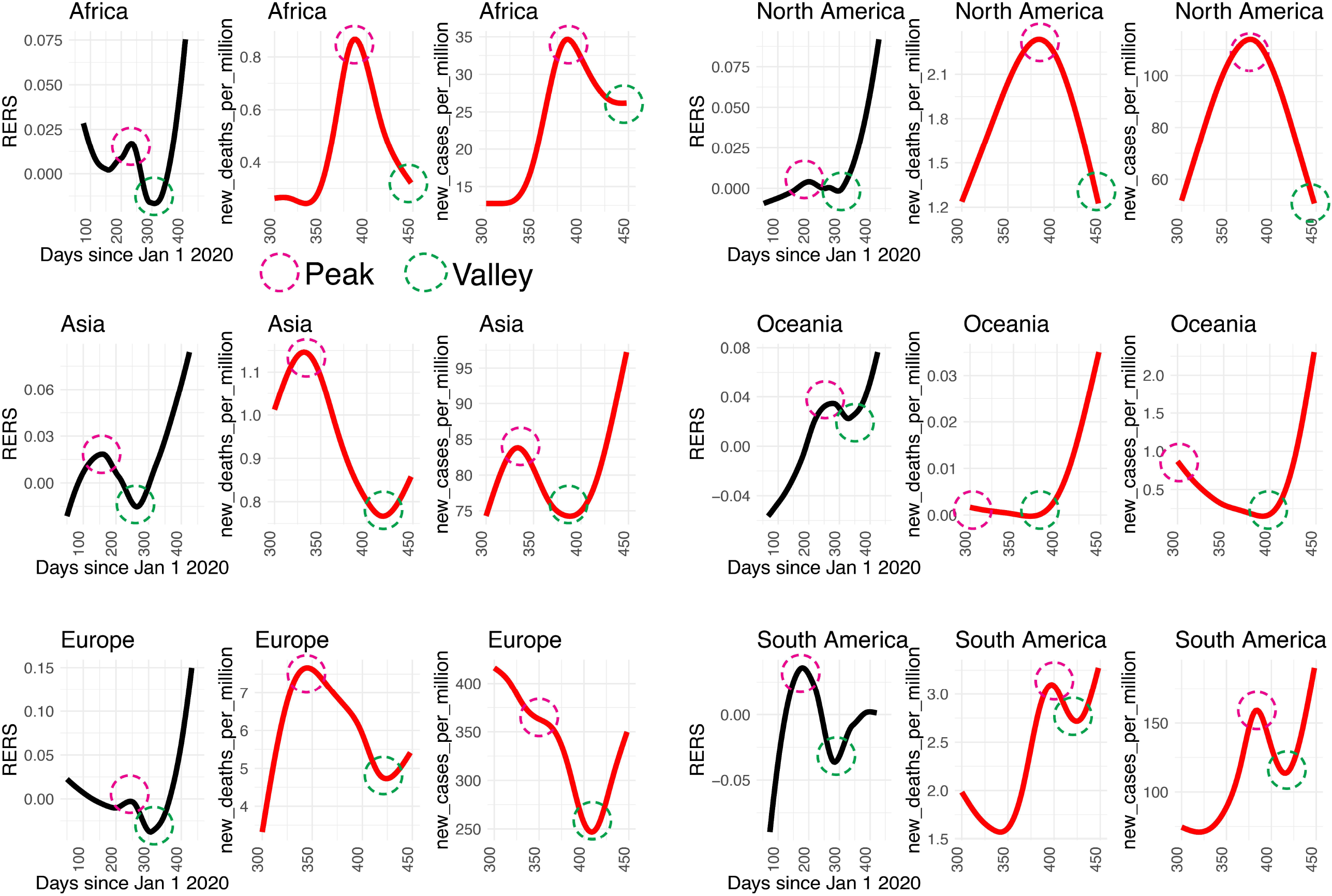
RERS and COVID-19 data fitting curves of each continent. The most recent waves in RERS (left, black, by LOWESS), new_deaths_per_million (middle, red, by Gamma Regression), and new_cases_per_million (right, red, by Gamma Regression) fitting curves of each continent. Red circle: peak, green circle: valley.

Based on the above results and analysis, we then predicted future pandemic trends based on current COVID-19 data with or without our RERS method (**Fig. 5**). Except in Oceania and South America, RERS trends were different from pure regression predictions in all the remaining continents, no matter using data of deaths (**Fig. 5A**) or cases (**Fig. 5B**). In Africa, Asia, Europe, and North America, both new cases and/or new deaths will continuously climb to a record-high point around July 2021.

**Figure 5:**
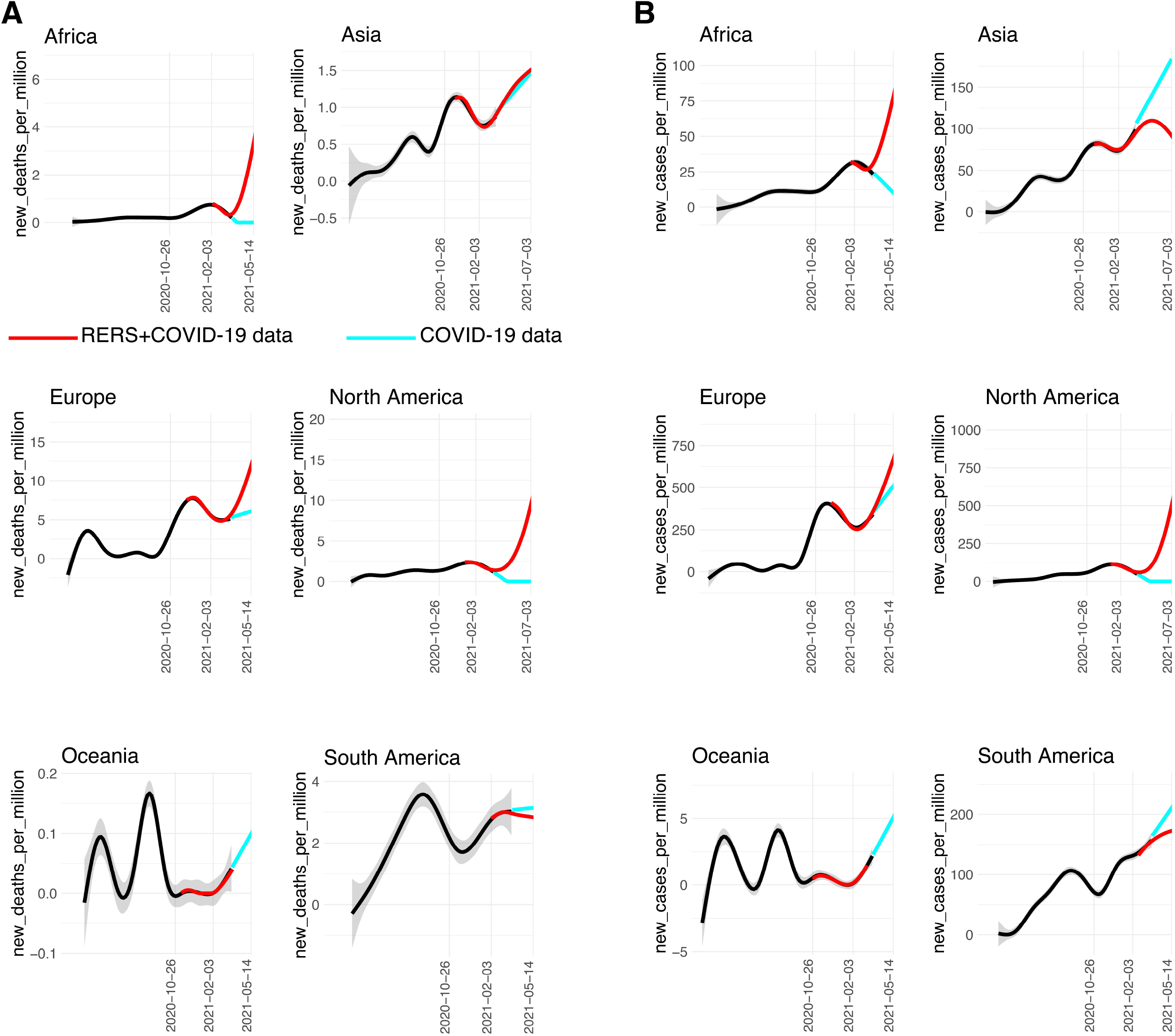
Prediction of future continental pandemic trends. (**A**) Predictions of each continent based on data of new_deaths_per_million with (red lines) or without (cyan lines) RERS. (**B**) Predictions of each continent based on data of new_cases_per_million with (red lines) or without (cyan lines) RERS. Gray shades: 95% confidence level interval, regression: Gamma Regression.

### Only Accelerated Spike Evolution Could develop New Variants

During the development of our bioinformatics approach, we observed that clade GR had both the highest genomic diversity percentage by distribution analysis (**Fig. 3A**) and fastest revolution by 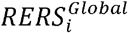 (**Fig. 3B**). Although it was the only exception in our finding that contributed most mutations yet wasn’t a small portion of SARS-CoV-2, it is still the truth that the latest clade GRY evolved out of it, found in the United Kingdom in December 2020 for the first time (*6, 12, 14*) and became the largest clade since February 2021.

Now that we predicted a future wave using 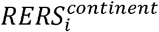, we would continue to explore whether these two clades could contribute to new variants which might potentially cause that wave. To do this, we separately analyzed MNPK and 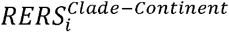 for SARS-CoV-2 of GR from December 17^th^ 2020 to March 15^th^ 2021 and of clade GRY from November 8^th^ 2020 to March 15^th^ 2021 from all continents. Through analysis, GR viruses generally showed a speeding-up mutation trend across all continents (**Fig. 6A**). By definition, MNPK larger than 2 denoted relatively high mutation and 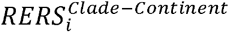 higher than 0.3 implied relatively high revolution (**Fig. 6B and 6C**). These highly revolutionary variants came from six countries of three continents, and included currently-found variants: P.1 (*18, 19*), B.1.1.28 (*19, 20*), B.1.1.317, and B.1.1.318 (*21*) (**Fig. 6D and 6E**). These variants of high MNPK and 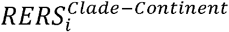 values positively showed a huge potential of clade GR to develop new variants in the future through continued fast revolution.

**Figure 6:**
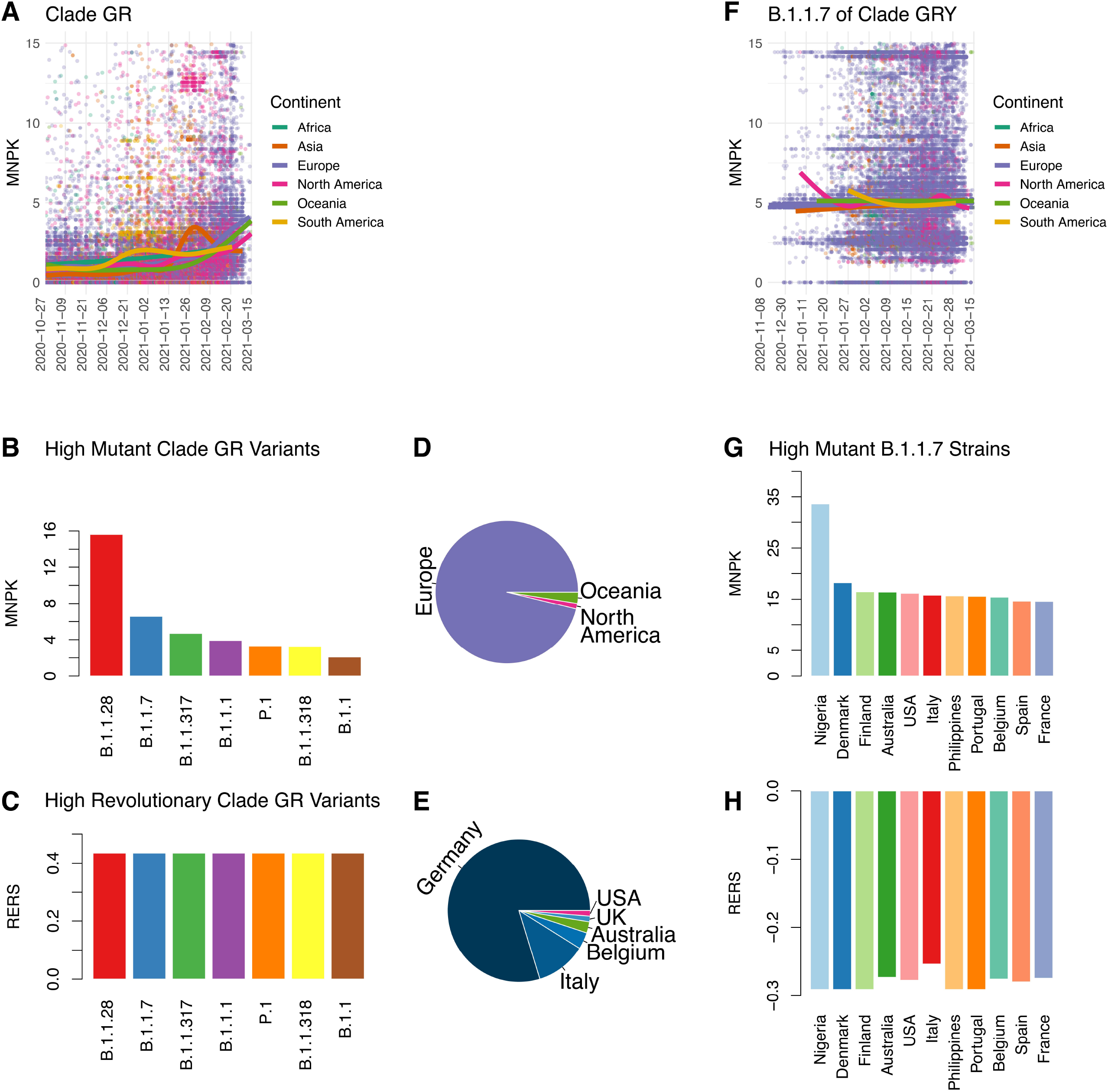
Combined MNPK and RERS analysis on clades GR and GRY. (**A**) MNPK of clade GR by continent. (**B**) High mutant GR variants were defined by MNPK values higher 2. (**C**) High revolutionary GR variants were defined as high mutant GR variants with RERS values higher than 0.3 in the meantime. (**D**) Continental origins of high revolutionary GR variants. (**E**) Corresponding country origins of the high revolutionary GR variants. (**F**) MNPK of B.1.1.7 of clade GRY by continent. (**G**) High mutant B.1.1.7 strains across countries were defined as those with MNPK values higher than 14.5. (**H**) RERS of these high mutant B.1.1.7 strains were all below zero.

On the contrary, B.1.1.7 in clade GRY showed a decelerated evolutionary trend in almost all continents (**Fig. 6F**), although all sampled strains from different countries had mutation rates higher than our threshold 14.5 (**Fig. 6G**). Together with the result that their 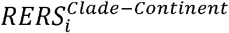 values were all below zero (**Fig. 6H**), new variants could not possibly develop out of it.

Conclusively, these results together suggested that sustained Spike revolution could be the most possible determinant of new variant’s emerging. This could be explained by the established selection theory that if mutations would benefit the survival of spreading of SARS-CoV-2, then they must be tested through a selection stage in a large population.

## Discussion

Tracking viral evolutionary pace is critical to better understand how emerged variants could impact global spreading. One famous example is that to understand how the HIV evolves from a next-generation sequencing angle is central to the development of appropriate drugs and vaccines (*22, 23*). We thus developed a bioinformatics approach to estimate the revolutionary rate over time by our defined metric, RERS. Distinct from the mutation rate evaluated by nucleotide substitutions/site/year for HIV, RERS focuses on changes or trends in the evolutionary process. Using RERS, we tracked Spike evolution by every 7 sampling days. Moreover, we developed an evolutionary-spreading model to integrate genomic Spike evolution data, COVID-19 data, related geographical indicators and related metadata. This model predicts a next pandemic peak by uniquely estimating the combined mutation and selection process.

Illustrated idea of our model is based on the combined mutation and selection theory in evolutionary biology (**Fig. 7)**. We found that the new variants carrying advantageous mutations could not be instantaneously dominant but has to experience a selection in a large population for 1 to 6 months. This course varies in the continents, indicating its relations with other factors, such as isolation policy, population size, and control policy (masking, distancing, and travel constriction). This may be also related to the transmissibility of the emerging variants (*6*).

**Figure 7:**
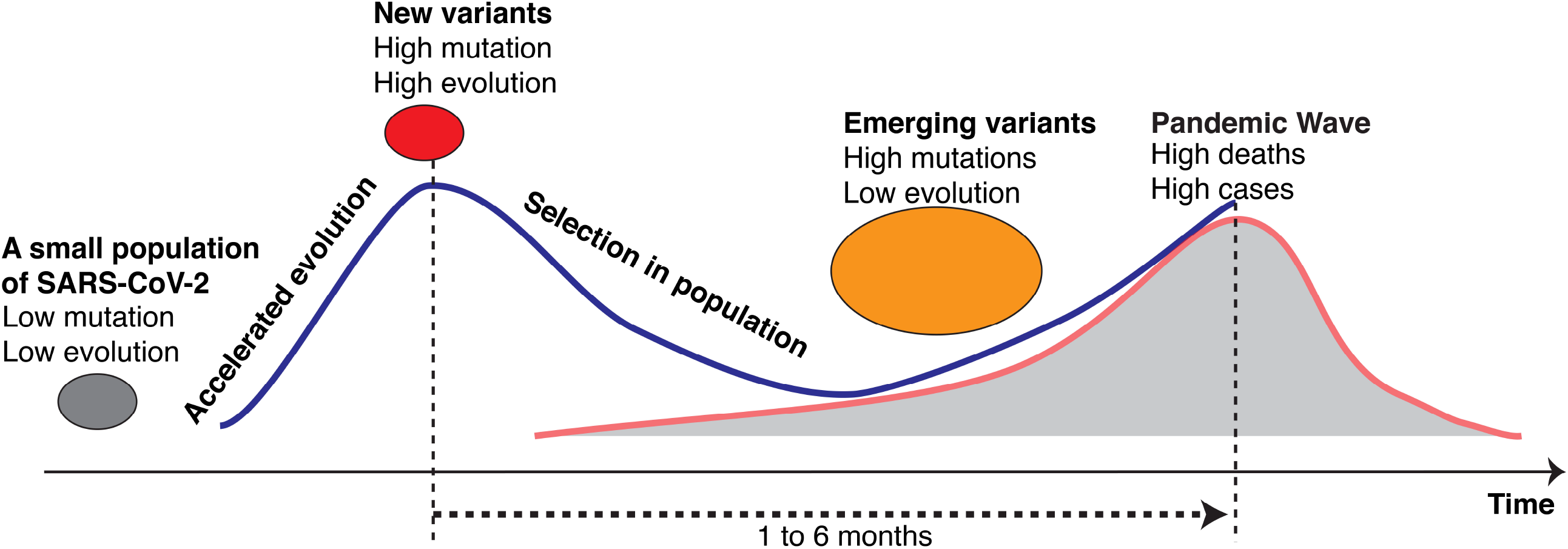
Illustrated revolution-selection process in COVID-19 pandemic. Increased Spike mutation happened first in a small population of SARS-CoV-2 (gray oval) then drove accelerated Spike evolution until its peak where new variants occurred with high mutation and revolution rates (red oval). Following a selection course of one to six months in large population, these variants (orange oval) on one hand became less evolutionary but on the other hand would lead to a pandemic wave of cases and deaths. Along with the increasing of infection and mortality, evolution went high again to possibly repeat this process in the future. Blue line: RERS fitting curve. Red line: COVID-19 data fitting curve. Gray area: cases and/or deaths number per day.

Distinct from existing studies only using the missense mutations at the protein level, we considered all types of mutations for the Spike genomic variations. The goal of our analysis is to estimate to what extent the virus mutates its genome, and to define an accurate mutation rate in terms of genomic single nucleotide variations and indels instead of only examining changes to amino acids. The necessity for including all mutations is also supported by recent discoveries for the genomic mutations in the C terminal of S protein in lineages B.1.1.7 and P.1, which are of unknown significance in associating with S protein functions (*12*). Our results confirmed that the virus has a low mutation rate normally but high dynamics when the virus reaches its limit because of the selective pressure. According to MNPK and RERS analysis, B.1.1.7 is the result of dramatically increased evolutionary rate (about 3-fold increased from October 2020 to March 2021) and taking about 6 months from its emergence to the current prevalent spread by a high selection. The emerged variants could be sponsored by an accelerated evolution during which some viruses, not necessarily the largest clade but could be relatively smaller ones, acquired the highest mutation rate.

We also analyzed MNPKs in different gender and age groups, found no difference among genders (**Fig. S4**). However, the largest mutations contributed by clades GH, GR, and GRY existed in people of 20-40 and 40-60 age groups compared to others (**Fig. S5**). This could explain why B.1.1.7 surges have shifted to younger population (*24*).

Genomic surveillance plays a key role in monitoring new variants and understanding the evolutionary spread (*25, 26*). Our model has revealed that the mutation and selection are critical to Spike evolution. Through gaining new mutations and surviving from the high selection pressure, current variants had the ability to spread more quickly in population (*3, 6, 21*), evade detection by specific diagnostic tests (*27*), and potentially evade natural or vaccine-induced immunity (*8, 12, 28*). Thus, the model could answer the question why cases and deaths were still increasing while people have been receiving vaccines: SARS-CoV-2 has been experiencing a second evolutionary acceleration since October 2020 and current variants such as B.1.1.7 already succeed selection thus would possibly reach a peak around July 2021.

So far, we have been equipped with several approved vaccines or candidates under phase 3 (*24*). Though it is urgent to evaluate if they are effective enough to compete Spike evolution, our model cannot make such a prediction due to insufficient vaccine data by March 23^rd^, 2021 (**Fig. S6**) but this will be our future focus after integrating evolutionary rate, pandemic data, and vaccine data. Future predictions would not only be useful to predict the pandemic trend post vaccination but also to evaluate vaccine effects in the context of Spike evolution.

## Methods

### SARS-CoV-2 and COVID-19 pandemic data

We downloaded the 817,351genome sequences from 6 continents and 185 countries/regions from GISAID (https://www.gisaid.org) as of March 15^th^, 2021. We also downloaded the sample metadata from GISAID, including information of geographic location, clade, lineage, collection date, as well as age and gender of confirmed cases. To associate the evolutionary data with the pandemic data, we downloaded the COVID-19 data from the World Health Organization (WHO) Coronavirus (COVID-19) Dashboard and the Our World in Data (https://ourworldindata.org/) as of March 23^rd^, 2021. The COVID-19 data include new_cases, new_deaths, new_cases_per_million, new_deaths_per_million, and vaccine information, such as people_fully_vaccinated_per_hundred.

### Mutation Rate in the Spike Genomic Sequence

We aligned the genomic sequence to the Spike reference from WIV04-reference (*13*), using the BLAST tools (*5*). We constructed a genome database for the 817,351genome sequences of SARS-CoV-2, downloaded from GISAID. We used BLAST for nuclei (blastn) as the alignment tool to align the 817,351genome sequences to the WIV04-reference. The newest version of BLASTN was downloaded from the National Institutes of Health BLAST website (https://ftp.ncbi.nlm.nih.gov/blast/executables/blast+/LATEST/), which was ncbi-blast-2.11.0+. The parameters used in the alignment are as follows and other parameters were used as default, - perc_identity 90 -max_target_seqs 10000000 -outfmt “6 qseqid sseqid sallacc stitle pident ppos gaps gapopen mismatch length qstart qend sstart send evalue bitscore score” We implemented the large-scale high-performance computing using Google Cloud Computing.

We defined the mutation rate by normalizing the number of mismatches and indels to the length of the Spike genomic region and multiplying 1,000. We denote the number of mismatches and indels as *n*, and the length of the Spike genomic region as *N*, and the mutation rate at the genomic level is

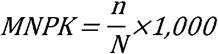

where MNPK represents Mutated Number Per Kilobases, evaluating the mutation rate of the Spike genomic region.

### Relative Evolutionary Rate of Spike Genomic Sequence (RERS)

To evaluate the mutation rate changes of the virus over time, we defined RERS to quantify the evolutionary rate. The definition of RERS is based on the MNPKs of the virus genomes over time. RERS is quantified by three steps: (1) using a window, e.g., 7 days, to group the genomes; (2) calculating average MNPK for each group of genomes; and (3) defining RERS for groups from second to last. RERS for a group of genomes is defined as the ratio between its average MNPK and that of the group prior to this group. Mathematically, we grouped the genomes by bins of days, generally 7 days, i.e., **G**_**1**_, **G**_**2**_,**…**, **Gk**. For *i* = *1, 2,…, g*, we calculated the mean MNPK for each group, i.e. *MNPK*_*1*_, *MNPK*_*2*_, **…**, *MNPK*_*k*_. Then, for *i* = *2*, **…**, *g*, we defined RERS as follows.

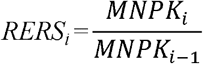

To show if the evolutionary rate is increased or decreased, we deducted RERS by 1.

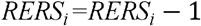

Thus, a positive value of RERS indicates a speed-up evolution whereas a negative RERS suggests a decreased pace of evolution.

### Statistical Regression of RERS and COVID-19 Data

To identify the trends and waves in the evolutionary rates and COVID-19 data, we applied the LOWESS regression (*29*) to the RERS data and the Gamma regression (*30*) to the COVID-19 data in the R ggplot2 package (https://ggplot2.tidyverse.org). The regression methods were automatically selected by the ggplot2 package. The regression methods from both LOWESS regression and Gamma regression are used to reveal trends and cycles in the data.

LOWESS (locally estimated scatterplot smoothing) regression combines the simplicity of the linear least squares regression with the flexibility of nonlinear regression. The model is based on a loss function *RSS*_*x*_*(A)* and a weight function *w(x)*. The traditional weight function for LOWESS is the tri-cube weight function *w*(*x*) = (1 − |*d*|^3^)^3^. The loss function is described as:

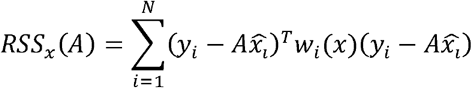

where 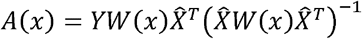.

Gamma regression is parameterized in terms of a shape parameter *α* =*k* and an inverse scale parameter *β* =1/*θ*, thus a Gamma distribution can be described as

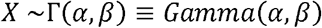

The probability density function in the shape-rate parameterization is

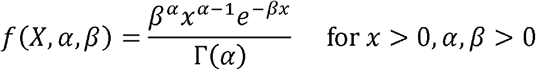

where Γ (*α*) - (*α* − 1)!.

### Correlation of RERS data with the COVID-19 data

To correlate the changes in the evolutionary rates and the corresponding changes in the pandemic data, we removed the course between the peak of RERS and the peak of the COVID-19 data and did data reshape based on the widths of the peaks in the RERS and the COVID-19 data (**Fig. S2**). We considered the COVID-19 pandemic data of new_deaths_per_million and new_cases_per_million. The relation between the evolutionary data (RERS) and the COVID-19 data is based on the mutation, selection, and spread processes associated with new variants, in which the incurred new cases and new deaths happen after the processes of mutation and selection of the new variants. We selected the latest peak and/or valley from a RERS distribution of a continent for association with the COVID-19 data. The RERS data from 200-450 days after Jan 1^st^, 2020 in Africa, Europe, South America, and Oceania, and the RERS data collected 100-450 days after Jan 1^st^, 2020 from Asia and North America were considered in the correlation analyses of the COVID-19 data.

After removing the course between the peak of RERS and the peak of the COVID-19 data, the peak of RERS can show consistent trends with that of the COVID-19 data. To find out the underlying association between these two types of data, we rescaled the peak of RERS by its width and that of the COVID-19 data. To accurately identify the widths of the peaks, we selected the summit point of the peak, *P*_*j*_, and the lowest point of the adjacent valley, *V*_*j*_ (**Fig. S2**). Thus, the width of a peak *j* is defined by *l*_*j*_ = *abs* (*V*_*j*_ − *P*_*j*_).

We reshaped the RERS peak by the peak width of RERS, *l*_*RERS,j*_ and that of the COVID-19 data, *l*_COVID-19,*j*_. The transformed data of RERS distribution were defined by the course between the RERS peak and the COVID-19 data peak, *C*_*RERS*→COVID-19,*j*_.

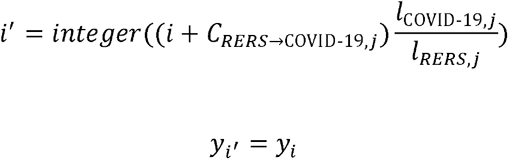

The correlations between the RERS and COVID-19 data were enabled by the reshaped distribution of RERS considering the peak widths. The statistical significances of the correlations were calculated by the reshaped distributions 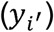 and the distribution of the COVID-19 data on the same days.

### Prediction of the upcoming trend of the COVID-19 data

We predicted the future trends of new cases and new deaths till July 3^rd^, 2021. Each prediction provides two trends: one boundary regressed by only COVID-19 data and another is the regression from the combined RERS and COVID-19 data.

The identified highly statistically significant correlation between RERS and the COVID-19 data provided the basis for our predictive model. The significant correlation suggests that the days used for mutation and selection may determine the spans of increase and decrease of the upcoming wave in the COVID-19 data that follows the mutation and selection processes.

For the prediction purpose, we not only reshaped the RERS data by removing the course between the peaks, *C*_*RERS*→COVID-19,*j*_ and rescaling the RERS data by the peak widths, but also reshape the RERS data by the summit values of the two peaks. We denoted the summit value of the RERS peak *j* as *s*_*RERS,j*_ and that of the COVID-19 data as *s*_COVID-19,*j*_. We rescaled the 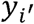 as follows.

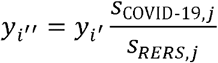

We used the Gamma regression to predict the future trends of the pandemic. Since our prediction is based on the course between the peaks of RERS and COVID-19 data (*C*_*RERS*→COVID-19,*j*_), our prediction can predict the trends up to days of the adjusted course (*i*′) from the day of the current peak summit of the COVID-19 data. The two boundaries for the prediction were determined by the regressions for the COVID-19 data only and the combined data (reshaped RERS+COVID-19 data).

### Statistical Analysis

Locally Estimated Scatterplot Smoothing, Pearson’s product–moment correlation coefficient, Welch two-sample *t*-tests (two-tailed), and Wilcoxon Rank-Sum test were implemented by R software (4.0.5).

## Data Availability

The SARS-CoV-2 genomic sequences are from GISAID (https://www.gisaid.org/).

## Acknowledgments

The authors of this study would like to acknowledge all the researchers who openly shared their genomic data on GenBank and GISAID. We also acknowledge the support of the Wake Forest Baptist Comprehensive Cancer Center Bioinformatics Shared Resource, supported by P30CA012197. The content is solely the responsibility of the authors and does not necessarily represent the official views of the National Cancer Institute. Funding: This work was supported by grants from Wake Forest Start-up funds (PI: Jin); Author contributions: G.J. was responsible for conception and design of the study and designed the computational analysis software and codes. Y.L and K.H were responsible for methods and hypothesis. G.X and N.Z took part in data analyses. Y.L, K.H, and G.J. wrote the paper.

