## Supplementary Figures and Legend for "Estimated Spike Evolution and Impact of Emerging SARS-CoV-2 Variants"

**Figure S1**

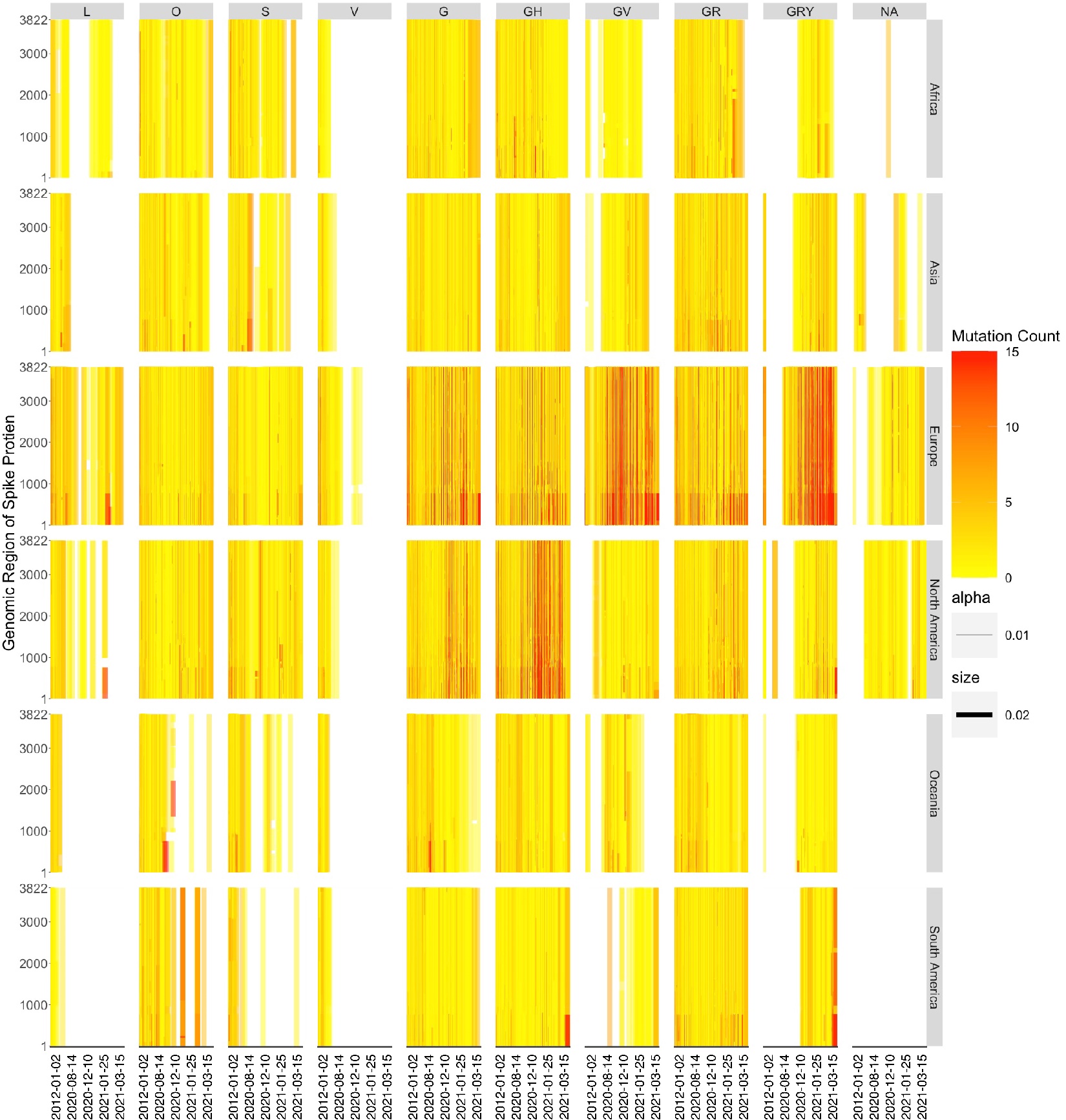

**Supplementary Figure 1: Mutation distribution across Spike genomic region over the global pandemic**. Mutation position in Spike coding region (vertical ordinate) are illustrated by clade (column) and continent (row) from January 1^st^ 2020 to March 15^th^ 2021 (horizontal ordinate). Each single line represents one SARS-CoV-2 genomic sequence. Columns are titled by CISAID clades and arranged from left to right according to their chronological order. Not assigned (NA) means SARS-CoV-2 sequences carrying mutations but are not classified into a new clade yet according to GISAID’s definition. Rows are titled by continents where SARS-CoV-2 sequences were collected and are arranged by alphabetical order from top to bottom.

**Figure S2**

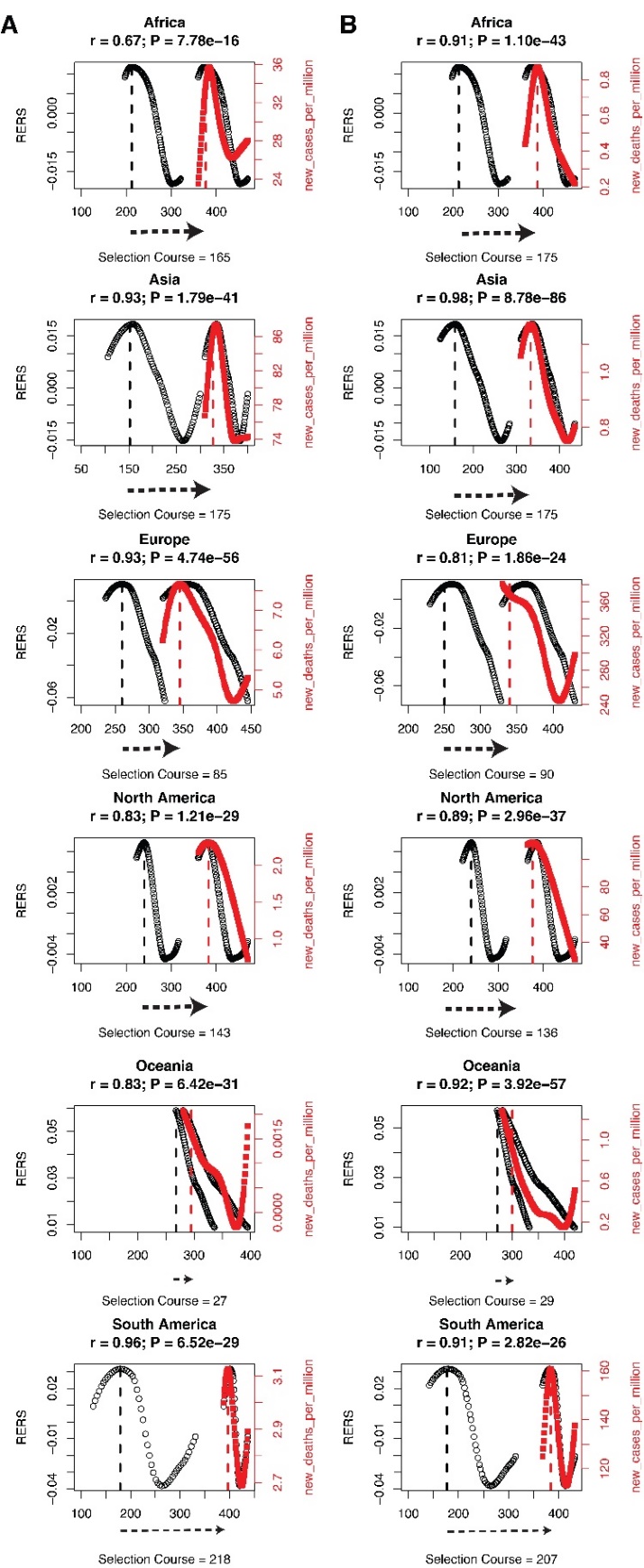

**Supplementary Figure 2: Correlation of peak-valley segments in RERS and COVID-19 data fitting curves.** Correlation of the most recent peak-valley segments in RERS and new_deaths_per_million curves (A) or in RERS and new_cases_per_million curves (B) by rescaling RERS segmental widths to fit COVID-19 segments. Lowest points were used when there were no valleys on fitting curves. Selection course was measure by days between the peaks of original RERS and COVID-19 fitting curves. Left black lines: original RERS segments associated with left y axes, right black lines: rescaled RERS segments associated with left y axes, red lines: COVID-19 segments associated with right y axes. Correlation Coefficient (*r*) and corresponding *P* values were calculated by the Pearson product–moment correlation coefficient.

**Figure S3**

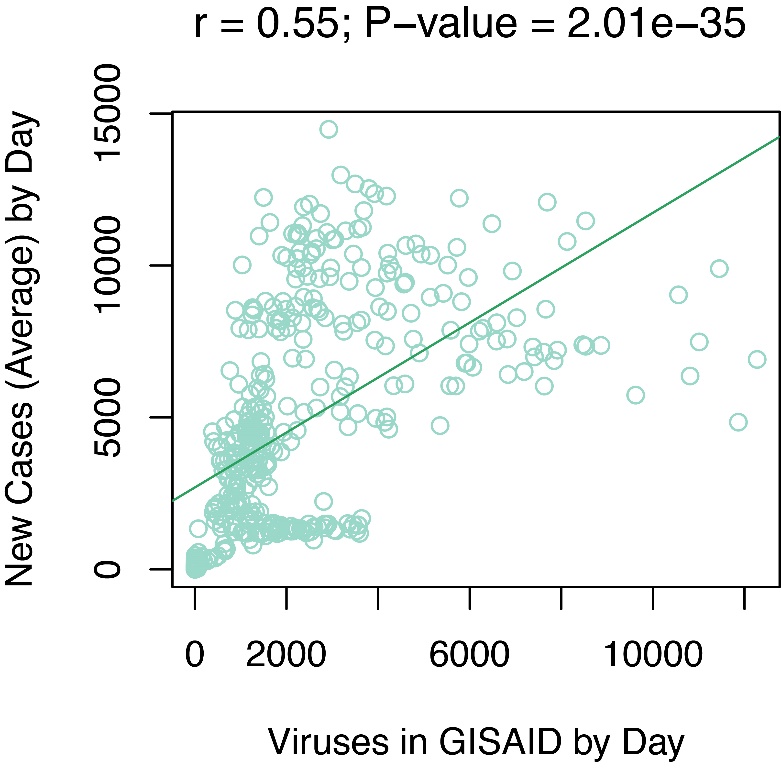

**Supplementary Figure 3: Correlation of collected SARS-CoV-2 genomic sample with new cases (average).** The value of new cases (Average) is calculated by dividing total case number by total source country numbers. Correlation was performed by Pearson’s product–moment correlation coefficient.

**Figure S4**

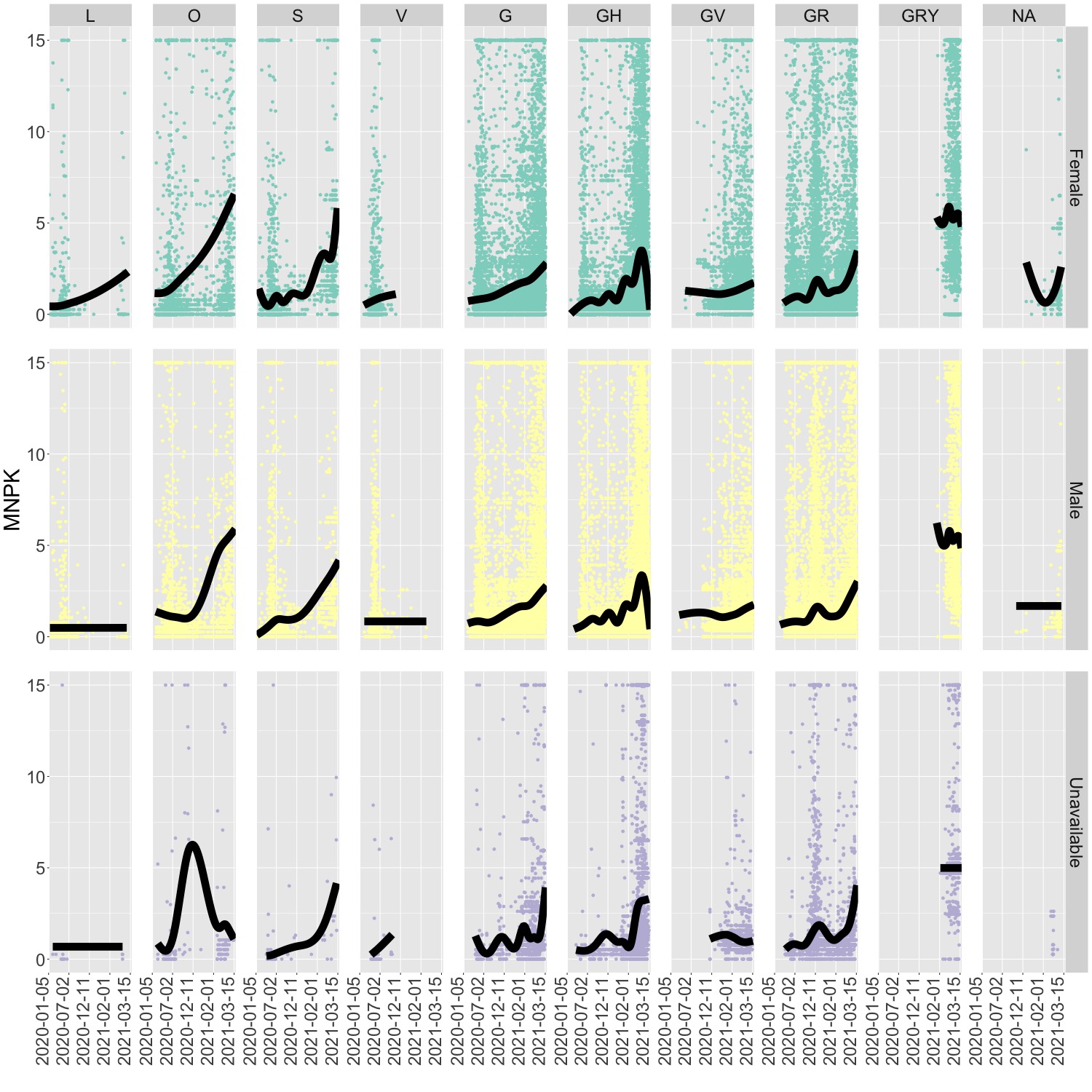

**Supplementary Figure 4: Mutation rates of the Spike genomic regions in all genders.** MNPK values (vertical ordinate) are illustrated by clade (column) and gender (row) from January 1^st^ 2020 to March 15^th^ 2021 (horizontal ordinate, segmented by every 100 sampling days). Columns are titled by CISAID clades and arranged from left to right according to their chronological order. NA as a column title means SARS-CoV-2 sequences carrying mutations but cannot be classified into a new clade yet according to GISAID’s definition. Rows are titled by genders from whom SARS-CoV-2 sequences were collected and are arranged from top to bottom by alphabetical order. Unavailable as a row title means no definite gender information from GISAID for some sequences. Black lines: fitting curves generated by Gamma Regression. Gray shades: 95% confidence level interval.

**Figure S5**

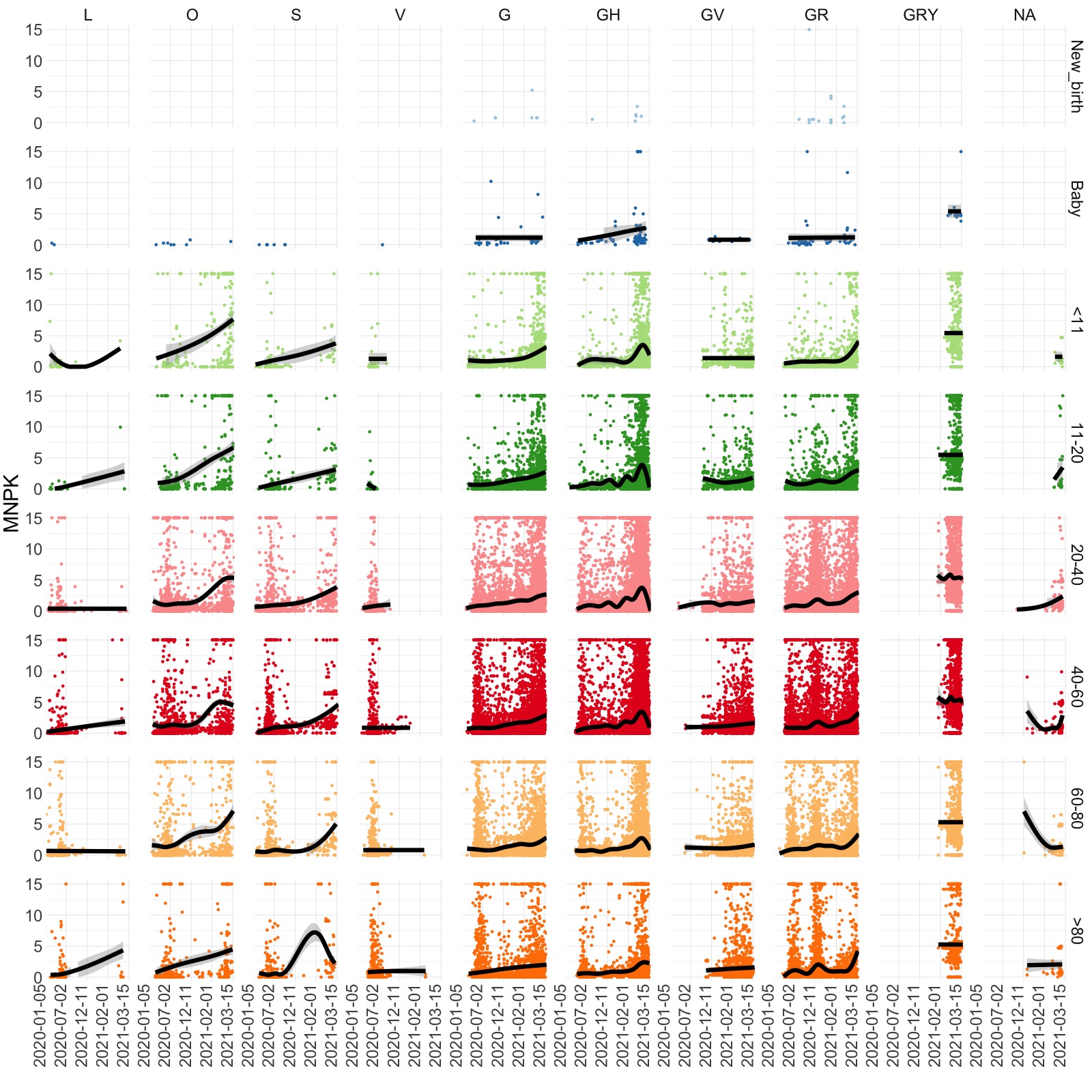

**Supplementary Figure 5: Mutation rate of Spike genomic regions in all age groups.** MNPK values (vertical ordinate) are illustrated by clade (column) and age group (row) from January 1^st^ 2020 to March 15^th^ 2021 (horizontal ordinate). Each single dot represents a mutation rate of one SARS-CoV-2 sequence calculated after blasting. Columns are titled by CISAID clades and arranged from left to right according to their chronological order. NA means SARS-CoV-2 sequences carrying mutations but are not classified into a new clade yet according to GISAID’s definition. Rows are titled by age groups from which SARS-CoV-2 sequences were collected and are arranged from top to bottom by an increasing order. New_birth: less than one month after birth. Baby: younger than one year old. Black lines: fitting curves generated by Gamma Regression. Gray shades: 95% confidence level interval.

**Figure S6**

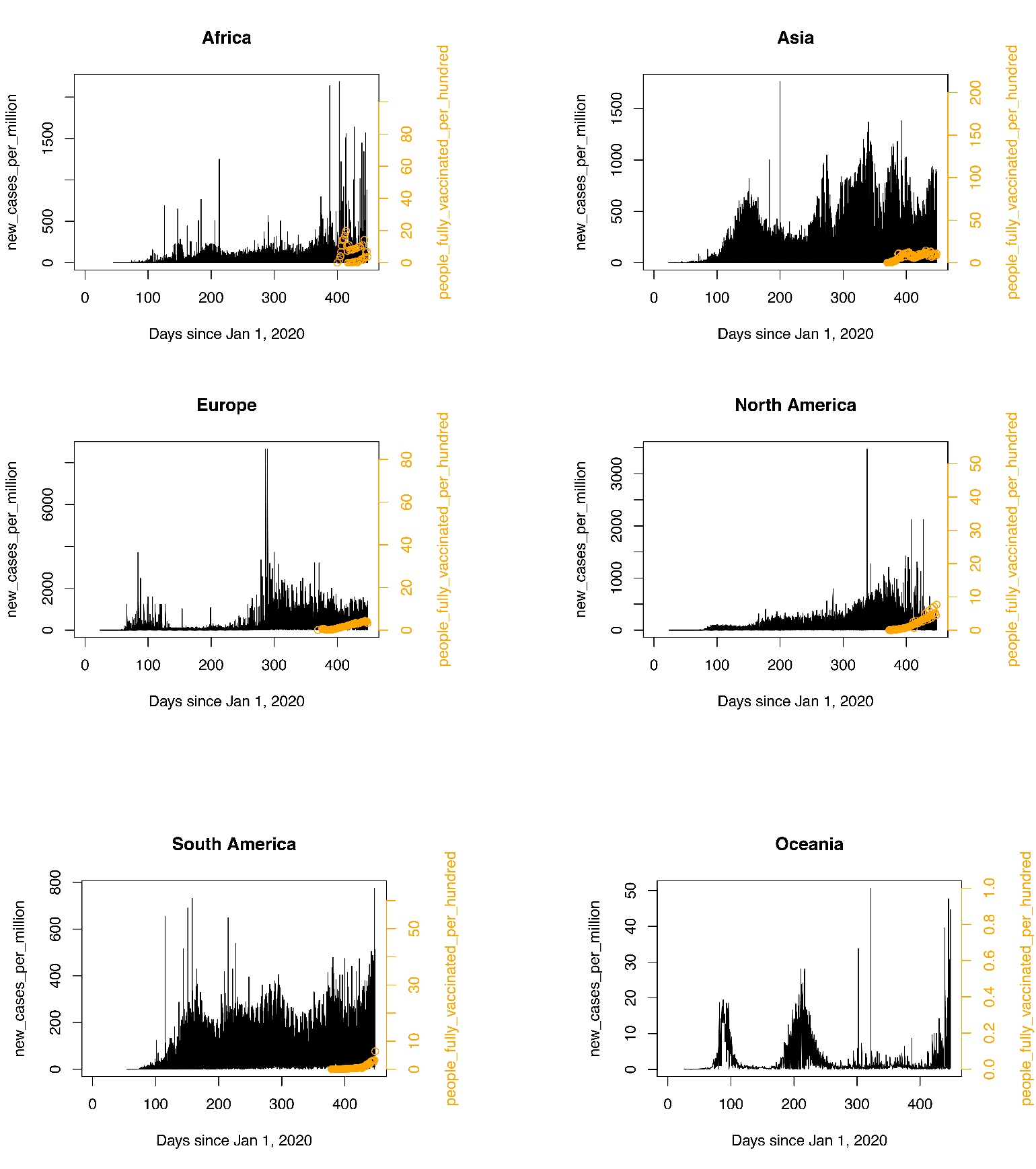

**Supplementary Figure 6: Available vaccine data compared with new cases by March 17, 2021 in each continent.** Black lines are showing new case numbers per day per continent since January 1^st^ 2020, which are associated with left *y* axis. Yellow circles are ratios of fully vaccinated people per hundred and associated with right *y* axis in each continent.
